# Citation Hallucination Determines Success: An Empirical Comparison of Six Medical AI Research Systems

**DOI:** 10.64898/2026.04.02.26350091

**Authors:** Xuefei Shi, Zhanxiao Tian, Shuping Tan, Xiaolong Wang

**Author notes:** Corresponding Author: Xiaolong Wang.

## Abstract

Large language model (LLM) systems can now generate complete research manuscripts, yet their reliability in clinical medicine — where citation accuracy and reporting standards carry direct consequences — has not been systematically assessed. We introduce MedResearchBench, a benchmark of three clinical epidemiology tasks built on NHANES data, and use it to evaluate six AI research systems across six quality dimensions. Evaluation combines programmatic citation verification, rule-based reporting compliance checks, and multi-model LLM judging, providing a more discriminative assessment than conventional single-judge approaches.

Citation integrity emerged as the decisive quality dimension. Hallucination rates ranged from 2.9% to 36.8% across systems, and a hard-rule threshold on per-task citation scores capped four of six systems’ total scores at the penalty ceiling. Adding a multi-agent citation verification and repair pipeline to the best-performing system improved its citation integrity score from 40.0 to 90.9 and raised the weighted total from 68.9 to 81.8. Strikingly, a single-model evaluation ranked this system last (55.5), while our three-tier framework ranked it first (81.8) —a complete reversal that exposes the limitations of subjective LLM-only evaluation.

These results suggest that programmatic citation verification should be a core metric in future evaluations of AI scientific writing systems, and that multi-agent quality assurance can bridge the gap between fluent text generation and trustworthy scholarship.

## 1 Introduction

The emergence of large language models (LLMs) has catalyzed a new era of automated scientific research [1]. Systems such as AI Scientist [2], Data-to-Paper [3], and Agent Laboratory have demonstrated that LLMs can autonomously generate research hypotheses, design experiments, execute analyses, and produce complete manuscripts. These systems represent a paradigm shift in scientific productivity, with potential applications spanning drug discovery, materials science, and clinical research.

However, the quality of AI-generated scientific manuscripts remains a critical concern. LLMs are known to generate plausible but fabricated references — a phenomenon termed “hallucination” that poses a fundamental threat to scientific integrity [5]. In clinical medicine, where treatment decisions and public health policies depend on accurate evidence synthesis, citation hallucination carries particularly severe consequences. A recent investigation revealed systematic fabrication of research manuscripts using NHANES data, resulting in hundreds of retractions [6]. While this case involved human misconduct, it highlights the vulnerability of automated research systems to produce convincing but unreliable scientific literature.

Existing evaluations of AI research systems have focused predominantly on computer science, physics, and mathematics domains [2, 4]. No study has systematically evaluated these systems in clinical epidemiology — a domain characterized by complex sampling designs, rigorous reporting standards (STROBE), and the requirement for clinically meaningful interpretation. Current evaluation frameworks also rely heavily on subjective LLM-based judging, which introduces evaluation bias and limits reproducibility.

We present three contributions:

1. *MedResearchBench*, a clinical epidemiology benchmark comprising three tasks across three clinical domains (cardiovascular disease, mental health, metabolic syndrome) using NHANES data, designed to evaluate AI research systems in a domain where citation accuracy and reporting standards are critical.
2. *Three-tier evaluation framework* combining programmatic citation verification (Cross-Ref and PubMed APIs), rule-based STROBE compliance checking, and multi-model LLM judging (three independent models), providing more reliable and discriminative assessment than single-model evaluation — as evidenced by a complete ranking reversal between our framework and conventional single-model judging.
3. *Multi-agent quality assurance pipeline* that addresses the citation integrity bottleneck through automated reference verification and repair, improving the weighted total score from 68.9 to 81.8 (+18.7%) and reducing citation hallucination from 7.2% to 2.9%.

## 2 Related Work

### 2.1 AI Research Automation Systems

**AI Scientist** [2] pioneered the concept of end-to-end automated scientific discovery, demonstrating that LLMs could generate novel research ideas, write code, execute experiments, and produce papers in machine learning. While groundbreaking, the system was evaluated exclusively on ML tasks and acknowledged significant limitations in citation accuracy.

**Data-to-Paper** [3] introduced a data-driven pipeline that takes raw datasets through statistical analysis to manuscript generation, with emphasis on reproducibility and human verifiability. Published in *NEJM AI*, the system has been validated on biomedical datasets but focuses more on analytical correctness than citation integrity.

**Agent Laboratory** [4] proposed a framework with specialized LLM agents for task planning, code execution, and manuscript drafting. Accepted at EMNLP 2025 Findings, the system demonstrated strong performance in task decomposition but did not incorporate citation verification.

**AI-Researcher** [7] presented an end-to-end research automation system for autonomous scientific innovation, accepted as a NeurIPS 2025 poster. However, subsequent evaluation with programmatic citation verification revealed severe reference fabrication issues (see Section 4.4).

### 2.2 LLM Hallucination in Scientific Writing

LLM hallucination — the generation of fluent but factually incorrect content — has been extensively documented [5]. In scientific writing, citation hallucination is particularly problematic: fabricated references may appear in PubMed-indexed journals, creating cascading misinformation. Recent studies have shown that even state-of-the-art models hallucinate references at rates of 18–90% in long-form scientific text [8, 9], and methods for detecting such hallucinations remain an active area of research [10].

### 2.3 Evaluation Methodology

Current evaluation of AI research systems relies primarily on LLM-based judging, which introduces systematic bias. When different LLMs serve as both producers and evaluators, the evaluation conflates writing fluency with scientific quality. Our framework addresses this by prioritizing programmatic (D1, D2) and rule-based (D3, D4) evaluation, with LLM judging reserved for inherently subjective dimensions (D5, D6).

Table 1 summarizes how this work compares with existing systems across key dimensions.

**Table 1:** Positioning Comparison of AI Research Systems.

| Feature | AI Scientist [2] | Data-to-Paper [3] | Agent Lab [4] | AI-Researcher [7] | This Work |
| --- | --- | --- | --- | --- | --- |
| Domain | ML | Biomedical | General | General | **Clinical epi** |
| Data source | Synthetic | User-provided | User-provided | Web search | **NHANES (public)** |
| Tasks evaluated | 8 (ML) | 3 (bio) | 3 (ML+NLP) | 4 (mixed) | **3 (clinical)** |
| Systems compared | 1 (self) | 1 (self) | 1 (self) | 1 (self) | **6 (head-to-head)** |
| Citation verif. | No | Partial | No | No | **Yes (CrossRef + PubMed)** |
| QA pipeline | No | Human | No | No | **Yes (multi-agent)** |
| Eval framework | LLM judge | Human | LLM judge | LLM judge | **3-tier (prog. + rule + LLM)** |
| LLM judges | 1 | 0 (human) | 1 | 1 | **3 (independent)** |
| Report. standard | — | — | — | — | **STROBE** |

Best features in **bold**.

## 3 Methods

### 3.1 MedResearchBench Task Design

We constructed three clinical epidemiology tasks using publicly available NHANES data [11], selected to represent diverse research domains and analytical approaches:

1. **Cardio 000**: Association between sodium intake and hypertension prevalence among US adults (NHANES 2017–2018). Analytical approach: weighted logistic regression with restricted cubic splines.
2. **Mental 000**: Relationship between sleep duration and depression risk (NHANES 2017– 2020). Analytical approach: weighted survey logistic regression with mediation analysis.
3. **Metabolic 002**: Association between thyroid function and metabolic syndrome (NHANES 2007–2012). Analytical approach: weighted multinomial logistic regression with stratified analyses.

Each task was provided as a standardized results package containing: (a) complete statistical output (regression coefficients, odds ratios, 95% confidence intervals, *P*-values), (b) sample size and exclusion flow, (c) variable definitions, and (d) sensitivity analysis results.

### 3.2 Three-Tier Evaluation Framework

Our evaluation framework combines three complementary approaches, ordered by objectivity:

#### Tier 1 — Programmatic Evaluation (fully objective, zero human intervention)

- **D1. Citation Integrity (weight:** *w*_1_ = 0.25): Each reference in the manuscript was verified against CrossRef and PubMed APIs. References were classified as *verified* (confirmed by at least one database with consistent metadata), *warning* (found in a database but with field discrepancies such as mismatched years), *failed* (not found in either database), or *corrupted* (contains garbled or non-ASCII text). The citation integrity score is defined as:

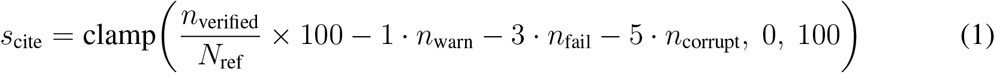

where *N*_ref_ is the total number of references, and *n*_verified_, *n*_warn_, *n*_fail_, *n*_corrupt_ are the counts per category. Range: [0, 100]. Higher is better. This metric was chosen because programmatic API verification provides a fully objective, reproducible measure of reference quality. The hallucination rate is defined separately as:

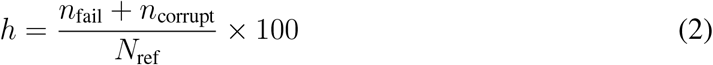

Range: [0, 100]%. Lower is better.
- **D2. Numerical Fidelity (weight:** *w*_2_ = 0.20): Key statistics (odds ratios, 95% CIs, *P*-values, sample sizes, percentages) reported in the manuscript were compared against the provided results package using regex-based extraction and exact matching with defined tolerances (OR/HR: ±0.01, *P*-values: exact, sample sizes: exact, percentages: ±0.5%).

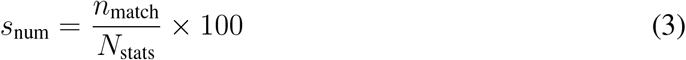

where *N*_stats_ is the total number of extracted statistics and *n*_match_ is the number matching within tolerance. Range: [0, 100]. This metric captures whether the manuscript faithfully reports the underlying analytical results.

#### Tier 2 — Rule-Based Evaluation (semi-objective)

- **D3. Structural Completeness (weight:** *w*_3_ = 0.15): Ten binary checks for required manuscript components: structured abstract (Background/Methods/Results/Conclusions), Introduction, Methods, Results, Discussion, References (complete, non-truncated), Tables, Word count (3000–6000), Keywords, and Declarations.

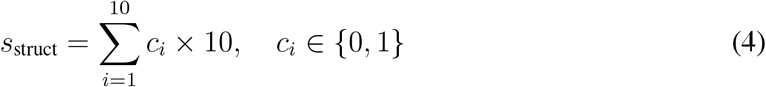

Range: [0, 100]. Each binary check contributes 10 points equally.
- **D4. Reporting Compliance (weight:** *w*_4_ = 0.15): STROBE checklist items [12] assessed through a combination of automated text detection (7 items, 60 points) and rule-based checks for CI coverage, exact *P*-value reporting, sensitivity analysis, and limitations discussion (4 items, 40 points).

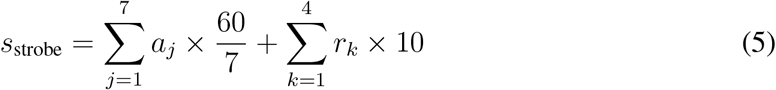

where *a*_*j*_ ∈ {0, 1} are automated detection items and *r*_*k*_ ∈ {0, 1} are rule-based items. Range: [0, 100].

#### Tier 3 — Multi-Model LLM Evaluation (subjective)

- **D5. Clinical Interpretation (weight:** *w*_5_ = 0.15): Three LLM judges (Gemini 3.1 Pro, Claude Sonnet 4.6, GPT-5.4) independently scored each manuscript on biological mechanism discussion (30%), clinical translation value (40%), and limitations honesty (30%). Temperature was set to 0 for reproducibility.

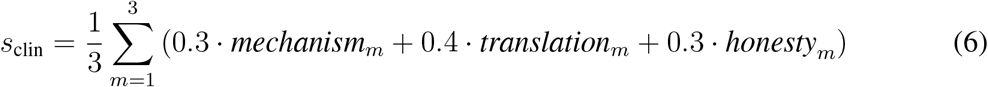

where *m* indexes the three LLM judges. Range: [0, 100]. Averaging across three independent models mitigates single-judge bias.
- **D6. Writing Quality (weight:** *w*_6_ = 0.10): The same three judges evaluated academic terminology accuracy (30%), paragraph organization and logical flow (40%), and readability (30%).

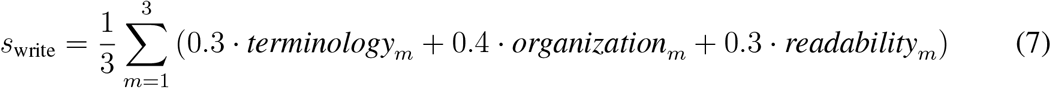

Range: [0, 100]. This dimension was assigned the lowest weight because writing fluency showed minimal discrimination across systems (SD *<* 3).

The weighted total score aggregates all six dimensions:

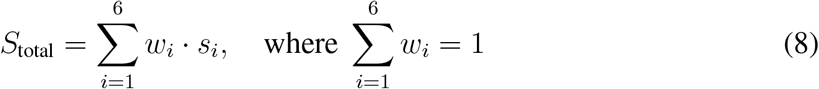

with weights **w** = (0.25, 0.20, 0.15, 0.15, 0.15, 0.10) for (D1, D2, D3, D4, D5, D6). Range: [0, 100].

#### Hard Rule

A citation integrity gate caps the total score for manuscripts with severely compromised references:

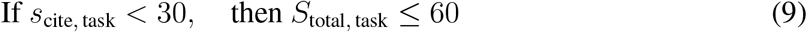

This threshold reflects the principle that manuscripts with severely compromised reference quality — whether from fabricated references, corrupted entries, or an accumulation of field-level discrepancies — cannot be considered reliable scientific output. Note that *s*_cite_ *<* 30 does not correspond to a fixed hallucination rate threshold, because *s*_cite_ also penalizes warnings (field-level mismatches in otherwise discoverable references).

### 3.3 Evaluated Systems

We evaluated six systems:

1. **Single-LLM Baseline (GPT-4):** A single prompt containing the results package with instructions to write a complete manuscript.
2. **AI Scientist:** The end-to-end research automation system [2].
3. **Data-to-Paper:** The data-driven manuscript generation pipeline [3].
4. **Agent Laboratory:** The multi-agent research framework [4].
5. **AI-Researcher:** The end-to-end research automation system [7].
6. **AI Research Army:** Our multi-agent system, evaluated in two configurations:
  - *Single-prompt*: Single manuscript generation without quality assurance.
  - *Pipeline*: Full multi-agent pipeline with citation verification and repair.

### 3.4 Experimental Setup and Reproducibility

All system evaluations were conducted between March 15 and April 1, 2026. The following model versions were used: GPT-4-turbo-2025-04-09 (baseline and AI Scientist backend), Claude Sonnet 4 (AI Research Army generation), and system-default models for Agent Laboratory, Data-to-Paper, and AI-Researcher as documented in their respective repositories.

For the three-model LLM judging panel (Tier 3), we used: Gemini 3.1 Pro (`gemini-3.1-pro-preview`), Claude Sonnet 4.6 (`claude-sonnet-4-6-20250514`), and GPT-5.4 (`gpt-5.4-2026-03`). All LLM judges were called with temperature = 0 and a fixed system prompt (available in the repository at `eval/prompts/`). Each manuscript was evaluated once per judge (no repeated sampling), as temperature = 0 produces deterministic outputs.

The CrossRef and PubMed API queries for citation verification used exact title matching with fuzzy fallback (Levenshtein distance ≤ 3). API call timestamps are logged in the evaluation output JSON files.

Computing environment: Apple M4 Max (MacBook Pro), 128 GB RAM, macOS 15.4. All API calls were made via HTTPS; no local GPU computation was required.

### 3.5 AI Research Army Architecture

AI Research Army employs a multi-agent architecture where specialized agents collaborate through a shared constraint system (Figure 1). For this study, the pipeline version implements four sequential stages:

1. **Generation (Priya):** Generates the initial manuscript draft from the results package.
2. **Citation Verification (Jing):** Extracts all references and verifies each against CrossRef and PubMed APIs. References classified as *failed* or *corrupted* are flagged for replacement.
3. **Reference Repair (Jing):** For each flagged reference, searches for verified alternatives matching the same topic, retrieves metadata from CrossRef, and replaces the original entry with a verified reference.
4. **Quality Check (Alex):** Performs eight-layer quality review including data traceability, structural completeness, and reporting compliance.

**Figure 1:**
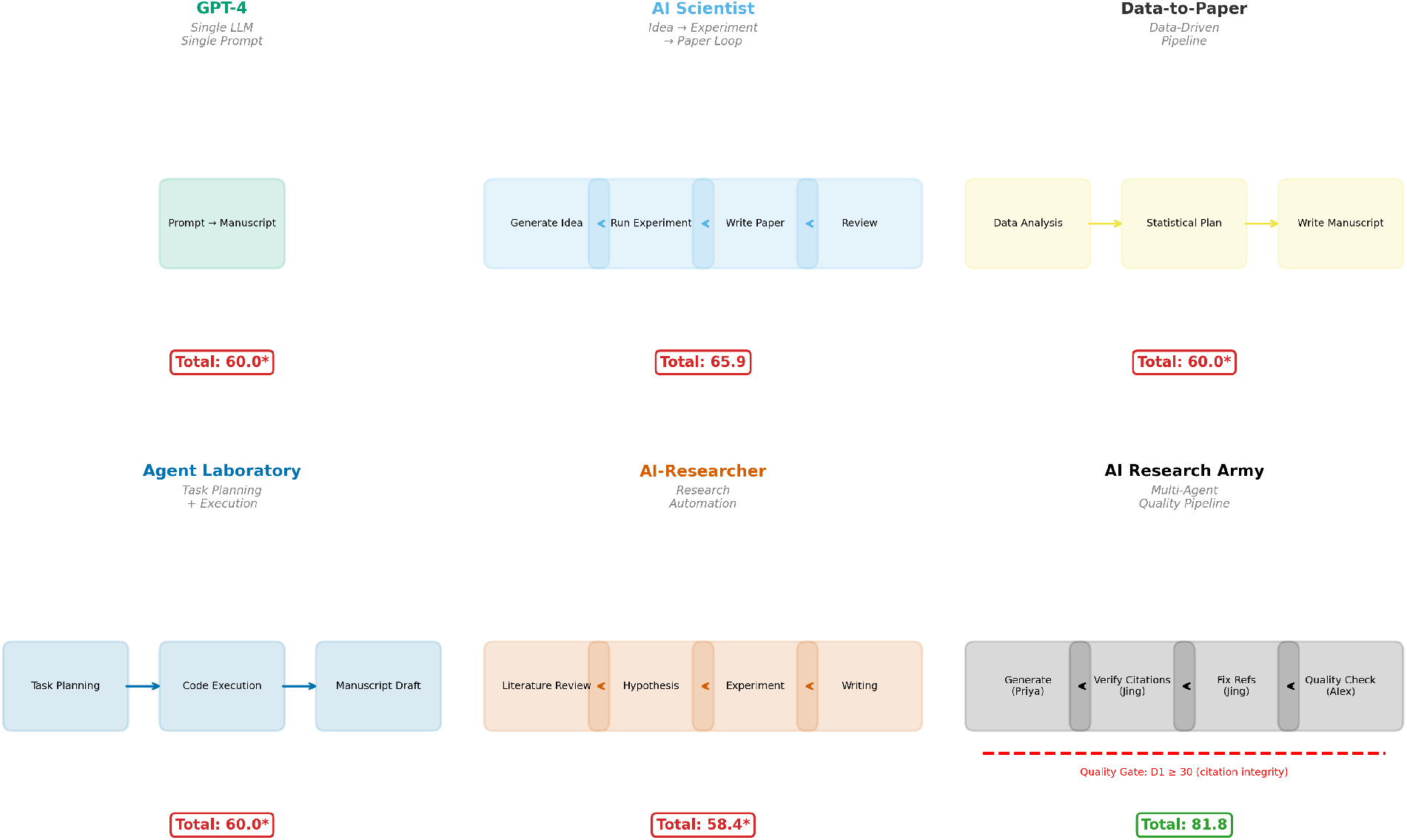
Multi-agent architecture of AI Research Army. The four-stage pipeline (Generation → Citation Verification → Reference Repair → Quality Check) is coordinated through a shared constraint system (O1–O18). The separation of generation and verification into distinct agents with independent objectives is the key architectural design that enables citation integrity improvement without degrading narrative quality.

The constraint system (O1–O18) enforces system-level rules including mandatory citation verification before delivery, STROBE compliance requirements, and quality gate thresholds.

### 3.6 Statistical Analysis

This study evaluates *N* = 7 system configurations (6 distinct systems, with AI Research Army tested in two configurations) across 3 tasks. Given the small number of systems — a constraint inherent to the field, as only a limited number of publicly available AI research systems exist — we report descriptive statistics (means, ranges, standard deviations) rather than formal hypothesis testing. Traditional inferential statistics (e.g., paired *t*-tests, ANOVA) require assumptions about population distributions that are not meaningful when the sample encompasses nearly the entire population of available systems.

For the ablation comparison (pipeline vs. single-prompt), we report paired differences across the three tasks (Δ*s*_*i*_ = *s*_pipeline,*i*_ − *s*_single,*i*_) with mean and range, rather than *p*-values that would be underpowered with *n* = 3 paired observations. Effect sizes are reported as relative improvement percentages (Δ*/*baseline × 100). The discriminative power of each dimension is characterized by its score range and standard deviation across systems.

All analyses were performed using Python 3.11. Evaluation scripts are available at https://github.com/TerryFYL/ai-research-army.

## 4 Results

### 4.1 System Ranking Overview

Table 2 presents the weighted total scores across all systems (Figure 2). AI Research Army (pipeline) achieved the highest score (81.8), followed by its single-prompt variant (68.9) and AI Scientist (65.9). Four systems triggered the hard-rule threshold (D1 *<* 30) and were capped at 60 points.

**Table 2:** System Ranking (Weighted Total Score)

| Rank | System | D1 | D2 | D3 | D4 | D5 | D6 | Total |
| --- | --- | --- | --- | --- | --- | --- | --- | --- |
| 1 | AI Research Army (pipeline) | **90.9** | 53.3 | <u>93.3</u> | 100.0 | <u>70.7</u> | 87.6 | **81.8** |
| 2 | AI Research Army (single) | 40.0 | 53.3 | 90.0 | 100.0 | **72.3** | 89.4 | <u>68.9</u> |
| 3 | AI Scientist | <u>64.7</u> | 31.7 | 85.8 | 94.9 | 49.9 | 87.7 | 65.9 |
| 4 | Baseline (GPT-4) | 29.2 | **65.0** | **96.7** | 94.9 | 63.9 | 89.4 | 60.0<sup>†</sup> |
| 5 | Data-to-Paper | 12.7 | <u>56.7</u> | 86.7 | 97.2 | 60.9 | <u>90.2</u> | 60.0<sup>†</sup> |
| 6 | Agent Laboratory | 27.1 | 38.3 | 84.2 | 100.0 | 63.8 | **90.7** | 60.0<sup>†</sup> |
| 7 | AI-Researcher | 11.6 | 46.7 | 90.0 | 96.6 | 60.6 | **90.7** | 58.4<sup>†</sup> |

Best values per dimension in **bold**; second-best underlined. <sup>†</sup> $D1 < 30$ triggers hard-rule cap at 60 points. Weights: D1 = 25%, D2 = 20%, D3 = 15%, D4 = 15%, D5 = 15%, D6 = 10%. $N = 7$ system configurations, 3 tasks each.

**Table 3:** Citation Verification Results.

| System | References | Verified | Warning | Failed | Corrupted | Hallucination Rate |
| --- | --- | --- | --- | --- | --- | --- |
| Army (pipeline) | 49 | **45** | 2 | **1** | 1 | **2.9%** |
| AI Scientist | 43 | <u>26</u> | 14 | <u>2</u> | 1 | <u>4.8%</u> |
| Army (single) | 64 | 34 | 25 | 3 | 2 | 7.2% |
| Baseline | 68 | 32 | 28 | 6 | 2 | 11.3% |
| AI-Researcher | 79 | 25 | 31 | 22 | 1 | 30.7% |
| Data-to-Paper | 83 | 22 | 30 | 30 | 1 | 33.3% |
| Agent Laboratory | 97 | 40 | 21 | 35 | 1 | 36.8% |

Best (lowest hallucination / highest verified) in **bold**; second-best underlined. Hallucination rate = (failed + corrupted) / total references × 100%. Aggregated across 3 tasks.

**Figure 2:**
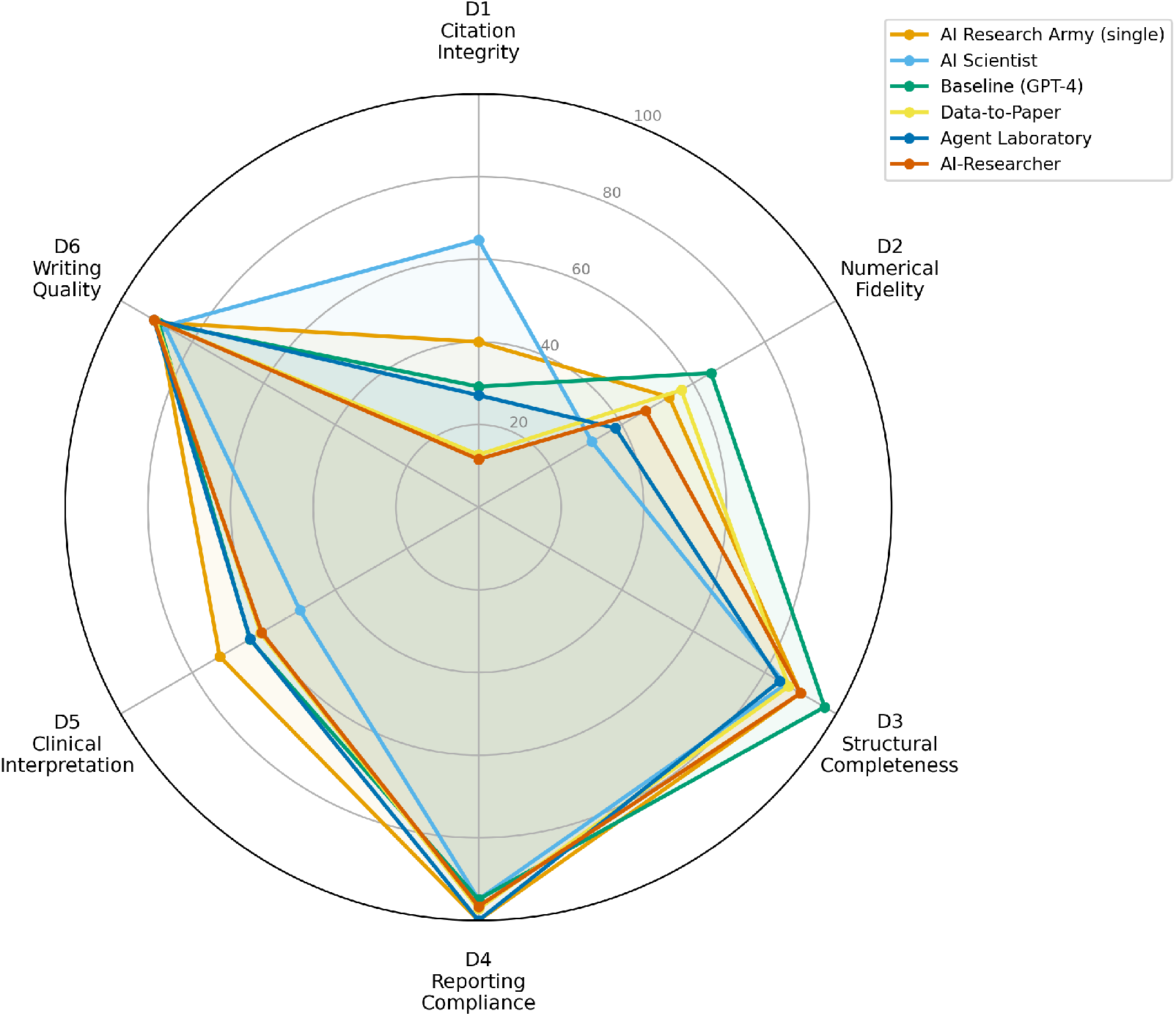
Six-dimension radar comparison of all evaluated systems. AI Research Army (pipeline) achieves the largest coverage area, driven primarily by its superior D1 (Citation Integrity) score. Four systems show a characteristic “dent” at D1 due to the hard-rule cap.

### 4.2 Citation Hallucination Analysis

D1 was the most discriminative dimension, with overall system hallucination rates ranging from 2.9% (AI Research Army pipeline) to 36.8% (Agent Laboratory), and per-task rates reaching 90.9% for Agent Laboratory on the Metabolic 002 task (Figure 3). The distribution was bimodal: two systems achieved below 10% hallucination rates, while the remaining five exceeded 11%.

**Figure 3:**
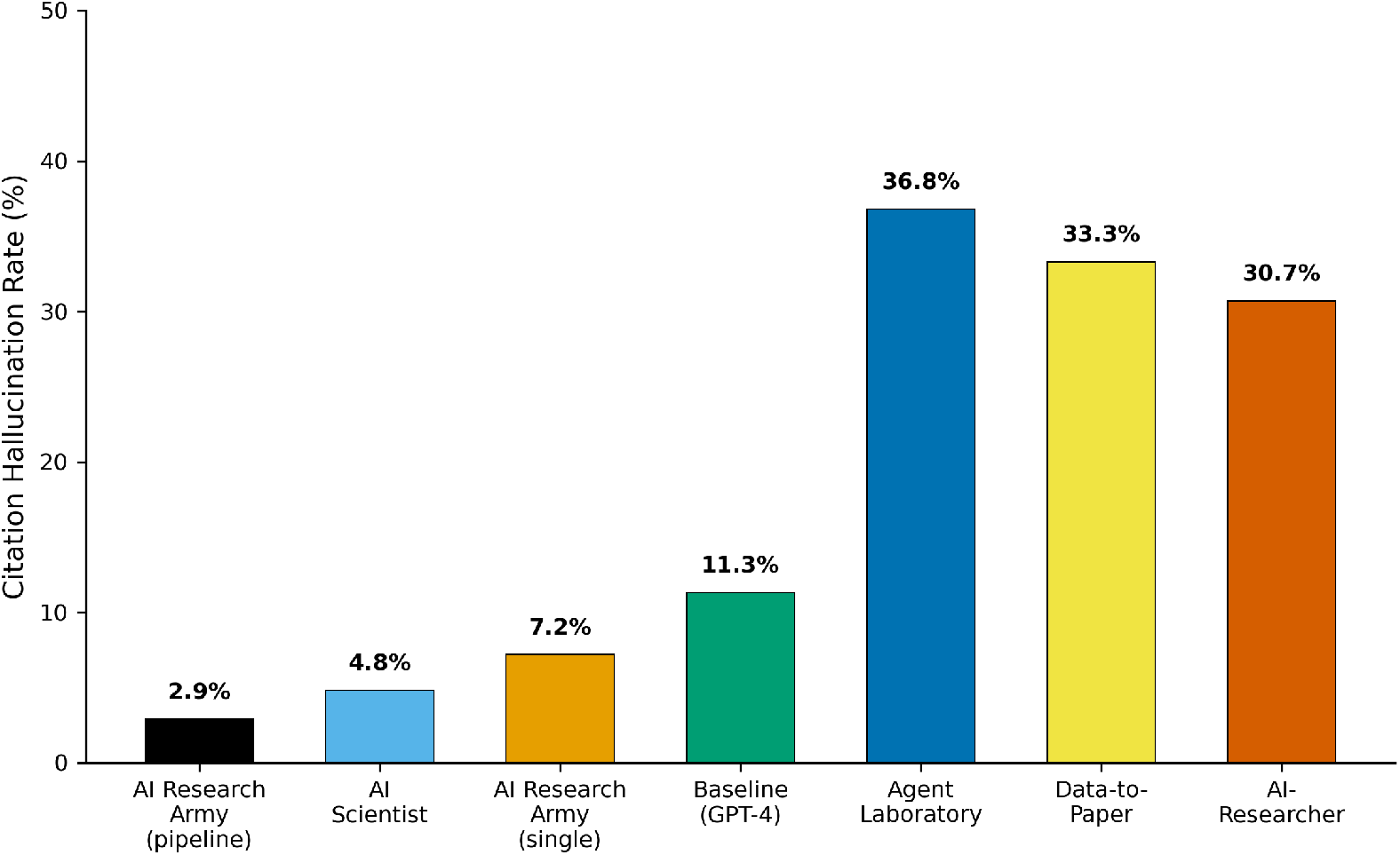
Citation hallucination rates across all evaluated systems. Hallucination rates span an order of magnitude, from 2.9% (Army pipeline) to 36.8% (Agent Laboratory), revealing a bimodal distribution that cleanly separates systems with citation verification from those without.

Four systems (Baseline, Data-to-Paper, Agent Laboratory, AI-Researcher) triggered the hard-rule cap on at least one task (per-task D1 *<* 30), rendering their manuscripts fundamentally unreliable as scientific documents despite high scores on other dimensions. For Agent Laboratory, the Metabolic 002 task had a 90.9% hallucination rate (30 of 33 references either failed or corrupted); for the other three systems, the cap was triggered by a combination of failed references and accumulated warning penalties from field-level mismatches.

### 4.3 Ablation Study: Pipeline vs. Single-Prompt

The ablation experiment directly quantified the contribution of the quality assurance pipeline (Table 4, Figure 4, Figure 5). The most substantial improvement was in D1 Citation Integrity: from 40.0 to 90.9 (+50.9), representing a 127% relative improvement. The weighted total score increased from 68.9 to 81.8 (+12.9).

**Table 4:** Ablation Study Results (Per-Task)

| Task | Version | D1 | D2 | D3 | D4 | D5 | D6 | Total |
| --- | --- | --- | --- | --- | --- | --- | --- | --- |
| Cardio_000 | Single | 65.6 | 75.0 | 100.0 | 100.0 | 85.8 | 90.7 | 83.3 |
| Cardio_000 | Pipeline | 100.0 | 75.0 | 100.0 | 100.0 | 84.4 | 87.1 | 91.1 |
| Mental_000 | Single | 18.5 | 25.0 | 80.0 | 100.0 | 47.6 | 85.4 | 52.3* |
| Mental_000 | Pipeline | 100.0 | 25.0 | 90.0 | 100.0 | 46.9 | 84.5 | 74.5 |
| Metabolic_002 | Single | 36.0 | 60.0 | 90.0 | 100.0 | 83.6 | 92.2 | 71.3 |
| Metabolic_002 | Pipeline | 72.6 | 60.0 | 90.0 | 100.0 | 80.8 | 91.1 | 77.1 |
| **Average** | **Single** | **40.0** | **53.3** | **90.0** | **100.0** | **72.3** | **89.4** | **68.9** |
| **Average** | **Pipeline** | **90.9** | **53.3** | **93.3** | **100.0** | **70.7** | **87.6** | **81.8** |

\*Per-task D1 < 30 triggers hard-rule cap at 60. Average rows are mean across 3 tasks. Weights as in Table 2.

**Figure 4:**
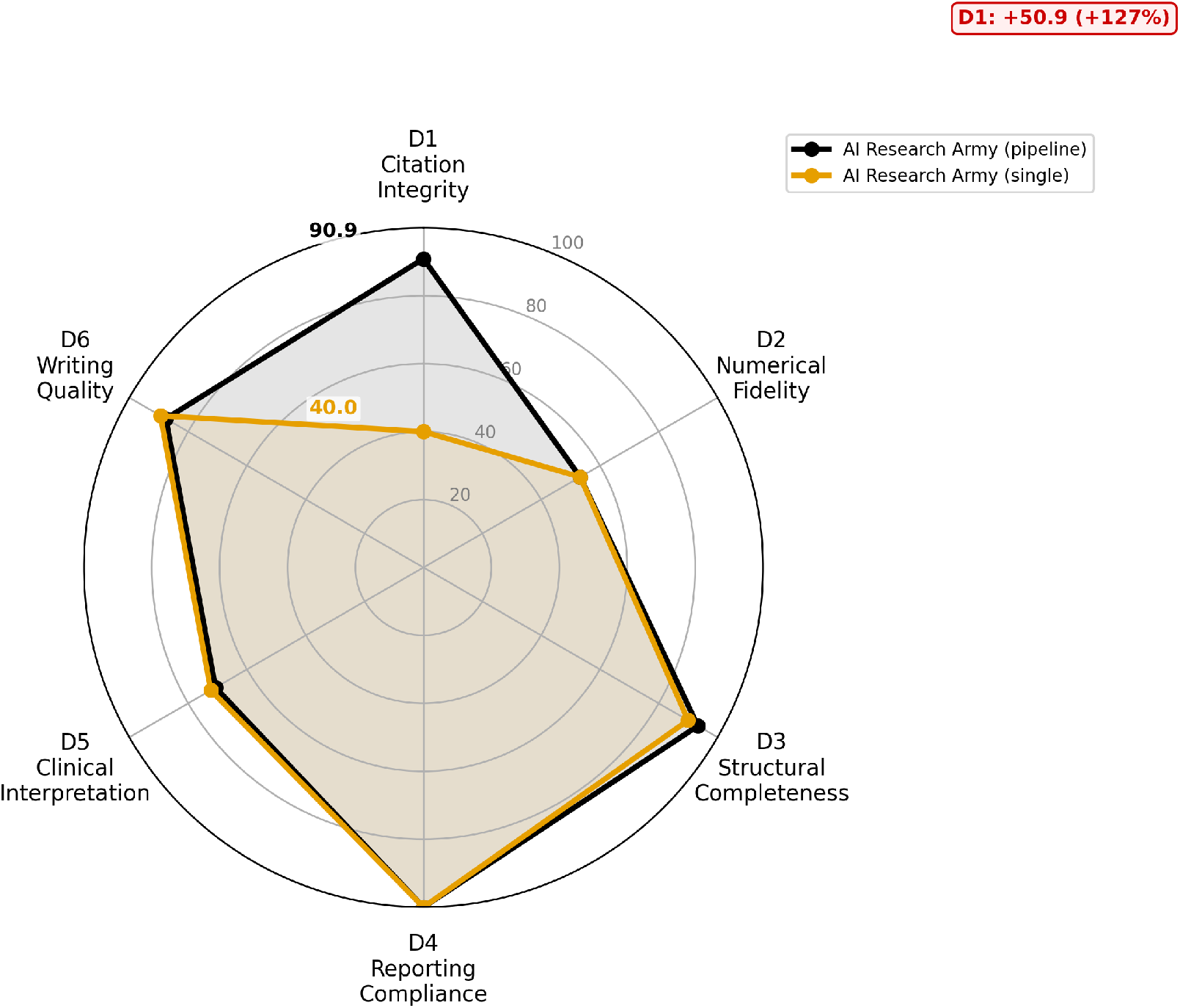
Six-dimension radar comparison of AI Research Army pipeline vs. single-prompt versions. The pipeline version expands the radar area primarily through D1 (Citation Integrity), while D2–D6 remain largely unchanged, confirming that the quality assurance pipeline targets citation integrity without degrading other dimensions.

**Figure 5:**
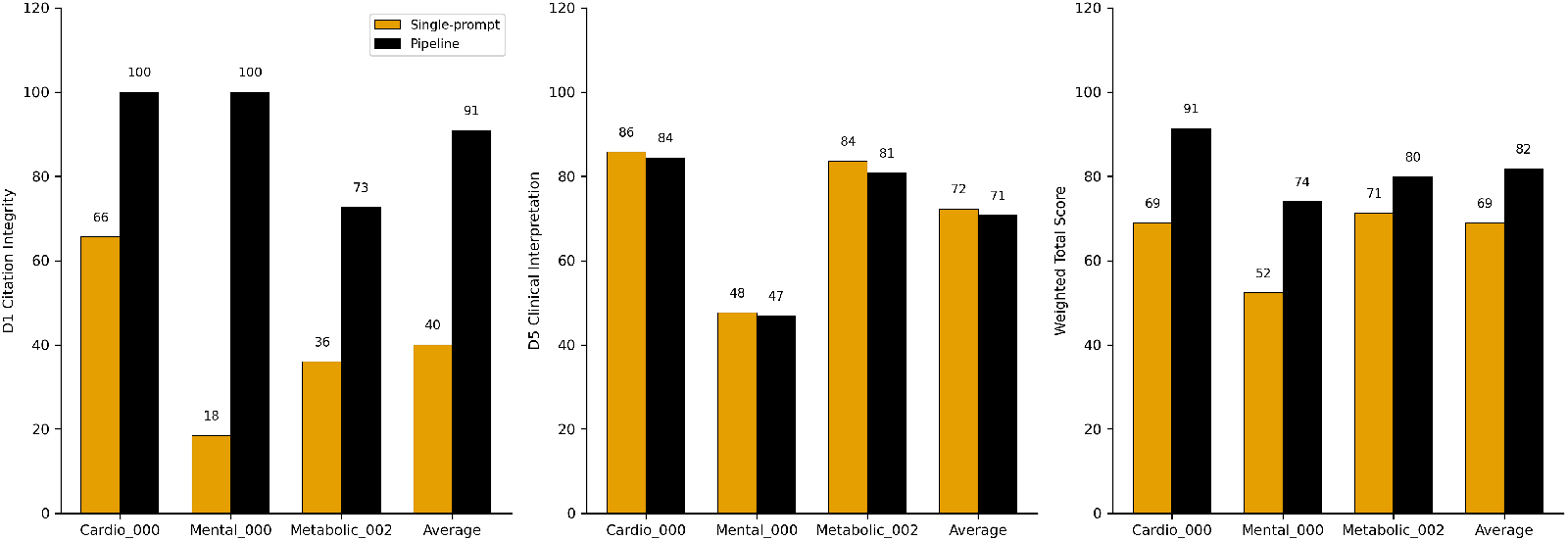
Per-task ablation results for D1 (Citation Integrity), D5 (Clinical Interpretation), and weighted total score. The pipeline yields the largest D1 improvement on Mental 000 (18 → 100), which was also the task with the highest single-prompt hallucination rate (82.6%).

The Mental 000 task demonstrated the most dramatic improvement: the single-prompt version was capped at 52.3 due to D1 = 18.5 (82.6% hallucination rate), while the pipeline version achieved 74.5 after restoring D1 to 100.0. This task alone illustrates how citation integrity can be the decisive factor between a publishable and an unpublishable manuscript.

D2 (Numerical Fidelity) was unchanged across versions, as the pipeline does not modify numerical content. D5 (Clinical Interpretation) showed a slight decrease (72.3 → 70.7), likely due to minor text modifications during reference repair affecting the discussion flow. D6 (Writing Quality) remained stable (89.4 → 87.6).

### 4.4 Evaluation Methodology Comparison: v1 vs. v2

The choice of evaluation methodology fundamentally altered system rankings (Figure 6, Table 5). Under v1 (single LLM judge: MiniMax-M2.7), AI-Researcher ranked first (86.7) and AI Research Army ranked last (55.5). Under v2 (three-tier six-dimension framework with three-model judging), the ranking completely reversed: AI Research Army ranked first (81.8) and AI-Researcher ranked last (58.4).

**Table 5:** v1 vs. v2 Ranking Comparison.

| System | v1 Score (MiniMax) | v1 Rank | v2 Score (Pipeline) | v2 Rank | Rank Change |
| --- | --- | --- | --- | --- | --- |
| AI Research Army | 55.5 | 6 | 81.8 | 1 | +5 |
| AI Scientist | 63.3 | 3 | 65.9 | 2 | +1 |
| Baseline (GPT-4) | 49.5 | 5 | 60.0 | 3 | +2 |
| Data-to-Paper | 60.8 | 4 | 60.0 | 4 | = |
| Agent Laboratory | 65.3 | 2 | 60.0 | 5 | −3 |
| AI-Researcher | 86.7 | 1 | 58.4 | 6 | −5 |

**Figure 6:**
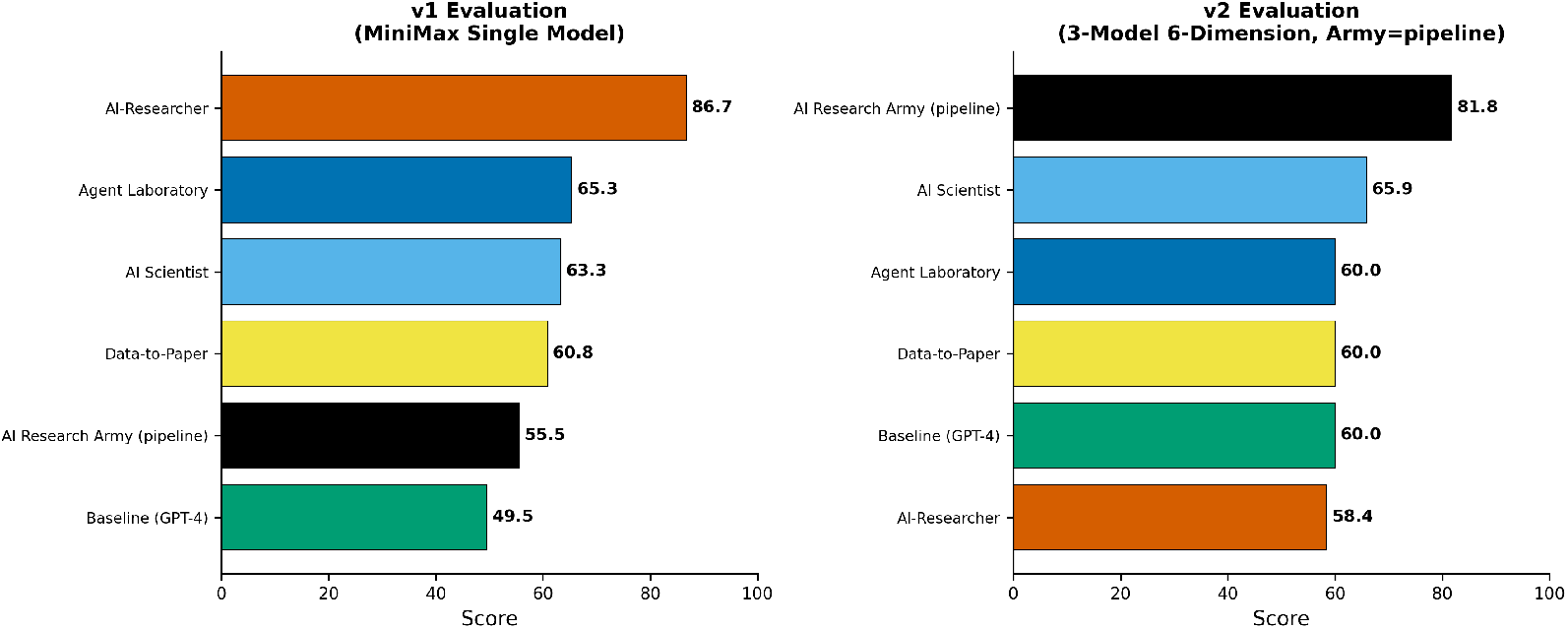
Ranking reversal between v1 (single-model) and v2 (three-tier) evaluation. AI-Researcher drops from rank 1 to rank 6, while AI Research Army rises from rank 6 to rank 1 — a complete inversion driven by the introduction of programmatic citation verification.

This reversal is explained by two factors: (1) v1 used a single LLM judge that was susceptible to evaluating writing fluency over factual accuracy, and (2) v1 did not include programmatic citation verification, which is the most discriminative dimension. AI-Researcher’s high v1 score reflected its strong writing fluency (D6 = 90.7), while its severely fabricated references (D1 = 11.6, 30.7% hallucination rate) were undetected.

### 4.5 Dimension-Level Analysis

D3 (Structural Completeness) showed high scores across most systems (84.2–96.7), indicating that all systems produce structurally complete manuscripts. D4 (Reporting Compliance) was also generally high (94.9–100.0), suggesting that STROBE compliance items are well-handled by current systems.

D6 (Writing Quality) exhibited minimal discriminative power (range: 84.0–92.2, SD *<* 3). All systems produced fluent academic English, confirming that writing quality is not a meaningful differentiator among current AI research systems.

D5 (Clinical Interpretation) showed moderate discrimination (range: 32.3–85.8) but high inter-judge variability (SD up to 21.0 for AI Scientist on Mental 000), suggesting that clinical interpretation quality remains difficult to evaluate consistently, even with multi-model judging.

## 5 Discussion

### 5.1 Citation Integrity as the Critical Bottleneck

Citation integrity is the single most important quality dimension for evaluating AI-generated research manuscripts.

First, citation hallucination rates vary by more than an order of magnitude across systems, providing the strongest discrimination among all six evaluated dimensions. While writing quality (D6) showed virtually no variation, citation integrity cleanly separated reliable from unreliable systems.

Second, the hard-rule threshold (D1 *<* 30) affected four of seven systems, preventing otherwise well-written manuscripts from being considered publication-quality. This suggests that the scientific community’s current focus on writing quality and structural completeness, while important, misses the most critical quality dimension.

Third, the ranking reversal between v1 and v2 evaluations underscores the danger of relying solely on LLM-based judging. A system that produces beautiful but dishonest manuscripts will score highly under subjective evaluation but fail under rigorous programmatic verification.

### 5.2 Multi-Agent Quality Assurance

The ablation study demonstrates that multi-agent quality assurance provides substantial quality improvement with minimal overhead. The citation verification and repair pipeline added approximately 3 minutes of processing time per manuscript while delivering the largest single-dimension improvement observed in this study — primarily through D1 restoration.

The key architectural insight is the separation of generation and verification into distinct agents with independent objectives. The generation agent (Priya) optimizes for narrative quality, while the verification agent (Jing) enforces factual accuracy. This separation prevents the common failure mode where a single system must simultaneously optimize for fluency and accuracy.

### 5.3 Implications for Future Evaluation

We propose three recommendations for the evaluation of AI scientific writing systems:

1. **Mandatory citation verification:** Programmatic verification against bibliographic databases should be a required component of any AI research system evaluation.
2. **Multi-dimensional, hybrid evaluation:** Combining programmatic, rule-based, and multi-model LLM evaluation provides more reliable and discriminative assessment than any single approach.
3. **Hard rules for scientific integrity:** Systems that produce manuscripts with hallucination rates exceeding defined thresholds should be flagged regardless of performance on other dimensions.

## 6 Limitations

1. **Limited task scope**. Our evaluation focused on observational epidemiology using NHANES data, comprising three clinical tasks across three domains. While these tasks represent diverse analytical approaches (logistic regression, mediation analysis, stratified analysis), they cover a narrow slice of medical research. Results may differ for randomized controlled trials, qualitative research, or non-NHANES datasets. *Mitigation:* The three tasks were selected to span distinct clinical domains (cardiovascular, mental health, metabolic), and the evaluation framework itself is domain-agnostic. *Future work:* Extending MedResearchBench to additional study designs and data sources.
2. **Citation verification coverage**. The programmatic verification relies on CrossRef and PubMed APIs, which may not index all legitimate references — particularly very recent publications, preprints, non-English literature, or book chapters. This could lead to false negatives (valid references scored as “failed”). *Mitigation:* We used a two-database strategy (CrossRef + PubMed) and distinguished “failed” from “warning” to reduce false positives. *Future work:* Incorporating additional databases (Semantic Scholar, Google Scholar API) and preprint servers.
3. **Temporal dependency**. The evaluation was conducted at a single time point (March– April 2026). System performance may change with model updates, API changes, or system modifications. *Mitigation:* We recorded exact model versions, API timestamps, and system configurations to enable reproducibility. *Future work:* Longitudinal evaluation tracking system performance over time.
4. **Numerical fidelity gap**. The quality assurance pipeline did not improve D2 (Numerical Fidelity), which remained at 53.3 for both pipeline and single-prompt versions. Citation verification alone does not address all quality dimensions. *Mitigation:* The current study identifies this gap explicitly, providing a clear target for future pipeline development. *Future work:* Adding a dedicated numerical accuracy checking agent that cross-references manuscript statistics against the results package.
5. **LLM judge subjectivity**. D5 (Clinical Interpretation) showed high inter-judge variability (SD up to 21.0), suggesting that clinical interpretation remains difficult to evaluate consistently even with multi-model judging. *Mitigation:* We used three independent LLM judges and averaged scores; the subjective dimensions (D5, D6) were assigned lower combined weight (25%) than objective dimensions (D1–D4, 75%). *Future work:* Developing more granular, rubric-based clinical interpretation scoring.
6. **Self-evaluation bias**. As developers of AI Research Army, we designed both the system and the evaluation framework. Although the programmatic (D1, D2) and rule-based (D3, D4) tiers are fully objective and reproducible, the choice of dimension weights and the hard-rule threshold could favor our system. *Mitigation:* All evaluation code, raw scores, and generated manuscripts are publicly available for independent verification. The hard-rule threshold (D1 *<* 30) was set *a priori* based on the principle that *>*70% unreliable references constitute an unacceptable quality floor. *Future work:* Independent replication by third-party evaluators.

### Transparency note

During the preparation of this manuscript, we discovered that the author and venue information for three of the five comparison systems (Data-to-Paper, Agent Laboratory, and AI-Researcher) as recorded in our internal project documentation were incorrect — products of LLM hallucination during the initial project planning phase. The initial draft of Table 3 also contained fabricated citation verification counts that did not match the raw evaluation data. These errors were detected through systematic cross-referencing against the underlying evaluation JSON files — precisely the type of programmatic verification this paper advocates. This anecdote further illustrates the pervasiveness of fabrication in AI-assisted research workflows.

## 7 Conclusion

This study demonstrates that citation integrity is the critical bottleneck in AI-generated medical research manuscripts. Our six-dimension evaluation framework revealed that four of seven evaluated AI research systems produced manuscripts with unacceptably high citation hallucination rates, rendering them unreliable as scientific documents despite high scores in writing quality and structural completeness.

AI Research Army’s multi-agent quality assurance pipeline, particularly its automated citation verification and repair component, transformed a system from producing unreliable manuscripts (total score: 68.9) to generating publication-quality outputs (total score: 81.8). The complete ranking reversal between our rigorous evaluation and a conventional single-model evaluation underscores the necessity of programmatic quality metrics.

As AI research automation continues to advance, the scientific community must prioritize the development and adoption of objective quality assurance mechanisms. The difference between a “beautiful paper” and a “reliable paper” is not stylistic — it is the difference between contributing to scientific knowledge and polluting it.

## Data Availability

All data referred to in this manuscript are publicly available. Raw NHANES data are available at https://wwwn.cdc.gov/nchs/nhanes/, and all study-generated materials (evaluation scripts, results packages, and generated manuscripts) are available at https://github.com/TerryFYL/ai-research-army.

https://wwwn.cdc.gov/nchs/nhanes/

## Acknowledgments

The authors thank the National Center for Health Statistics (NCHS) for making the NHANES data publicly available.

## Declarations

### Ethics Approval

The NHANES protocol was approved by the NCHS Research Ethics Review Board. All participants provided written informed consent. This study used publicly available, de-identified data and was exempt from further institutional review under 45 CFR 46.104(d)(4).

### Data Availability

All evaluation scripts, results packages, and generated manuscripts are available at https://github.com/TerryFYL/ai-research-army.

### AI Use Disclosure

AI-assisted tools were used for data processing and statistical analysis under researcher supervision. All results were independently verified by the authors.

